# Emergency admission plasma D-dimer and Prothrombin activity in acute ischemic stroke with large artery occlusion

**DOI:** 10.1101/2024.09.24.24314335

**Authors:** Shandong Jiang, Peizheng Guo, Cong Qian, Jun Yu, Liang Xu, Xu Li, Xianyi Chen, Fang Bing, Yuan Yuan, Zhongju Tan, Jing Xu, Jianru Li

**Author notes:** Corresponding authors: **Jing Xu,** Department of Neurosurgery, Second Affiliated Hospital, School of Medicine, Zhejiang University, Clinical Research Center for Neurological Diseases of Zhejiang Province, Hangzhou, 310009, China. **Jianru Li**, Department of Neurosurgery, Second Affiliated Hospital, School of Medicine, Zhejiang University, Clinical Research Center for Neurological Diseases of Zhejiang Province, Hangzhou, 310009, China. S.J. and P. G. contributed equally to this work.

## Abstract

**Background:** The coagulation system is intrinsically linked to pathological mechanism and progression of ischemic stroke. However, the role of preoperative coagulation function in determining the functional outcomes of acute ischemic stroke patients following large artery occlusion (AIS-LVO) has not been extensively evaluated in peer-reviewed literature.

**Methods:** We utilized logistic regression analyses, complemented by the construction of receiver operating characteristic (ROC) curves, to identify significant predictive factors for poor prognosis following EVT. Additionally, subgroup analyses were conducted to further assess the prognostic efficacy of coagulation function across different subgroups.

**Results:** A total of 607 patients were enrolled, with 335 (55.19%) experiencing an unfavorable outcome. Multivariate regression analysis identified preoperative D-dimer and PTA as independent predictors of 3-month prognosis. After adjusting for confounders, elevated preoperative D-dimer levels (≥715 mg/L), identified by cut-off value, were a significant predictor of poor prognosis, with 2.51-fold higher risk compared to the normal range. Conversely, elevated PTA levels (≥85.5%) were significantly and inversely associated with poor prognosis, indicating a reduced risk of 0.39 times. Furthermore, the combination of elevated D-dimer and reduced PTA demonstrated a synergistic effect, markedly increasing the risk of poor outcomes in AIS-LVO patients. Subgroup analyses revealed that failed recanalization, comorbid diabetes, and non-middle cerebral artery (MCA) occlusion significantly influence the predictive value of D-dimer and PTA for clinical outcomes.

**Conclusion:** Elevated admission D-dimer and reduced PTA levels are an independent predictor of poor prognosis in patients with AIS-LVO, and there is a synergistic interaction between the two variables.

## Introduction

Acute ischemic stroke (AIS) is the second most lethal and disabling disease worldwide, accounting for approximately 80% of all strokes. It severely impairs patients’ survival and quality of life, leading to significant economic and social consequences[1]. Clinically, about one-third of AIS patients are acute large vessel occlusions (AIS-LVO), which present with more severe clinical symptoms and higher rates of disability and mortality compared to non-large vessel occlusion strokes[2]. In 2015, five landmark randomized controlled trials (RCTs) established the superiority of endovascular thrombectomy (EVT) over intravenous thrombolysis (IVT) for the treatment of anterior circulation AIS-LVO[3]. EVT, including endovascular contact aspiration and stent retriever techniques, has become a cornerstone therapeutic approach for AIS-LVO, extending from anterior to posterior circulation cerebral infarctions[4]. Technological and material advancements have led to a marked increase in the recanalization rate among AIS-LVO patients, rising from 58%-65% in 2015[3]to an impressive 83.1% - 92%[5]. However, despite these improvements, the overall prognosis remains suboptimal, with a significant proportion of patients (47.6%-59.8%) failing to achieve a favorable outcome or functional independence post-EVT[6,7]. Therefore, it is important to explore the risk factors of unfavorable prognosis in patients with AIS-LVO after EVT, which may help guide treatment decisions and the expectations of patients and their families.

In clinical practice, certain clinical features have been identified as predictors of poor prognosis in AIS patients post-EVT, such as age[8], National Institutes of Health Stroke Scale (NIHSS) scores[9], hyperglycemia[10], dyslipidemia[11], and pass attempt exceeding three times during the procedure[12]. However, some of these indicators, like NIHSS or modified Rankin Scale (mRS) scores, are subject to variability due to differences in assessment across hospitals and individuals. Due to the clinical ubiquity and routine of preoperative testing, researchers has increasingly focused on the diagnostic and prognostic value of various auxiliary indices. These objective measures offer the potential for more standardized and reliable prognostic assessment, which is crucial for guiding treatment decisions and setting realistic expectations for patients and their families, ultimately aiming to improve patient outcomes following EVT[13,14].

From a pathophysiological perspective, AIS is characterized by dynamic changes in the coagulation system during its acute phase. The modulation of thrombin generation and the coagulation cascade has been a longstanding target for both the treatment and prevention of AIS[15]. Elevated brain thrombin levels, detected in the infarct area following ischemia, are attributed to the breakdown of the blood-brain barrier (BBB) and the entry and synthesis of prothrombin in the brain[16], playing a significant role in the brain’s pathological response to ischemic stroke. As part of the routine preoperative examination, coagulation function indicators, including D-dimer, international normalized ratio (INR), prothrombin time (PT), activated partial thromboplastin time (APTT), prothrombin activity (PTA), thrombin time (TT), and fibrinogen (FIB), provide insights into both the extrinsic and intrinsic coagulation pathways[17], which may be closely associated with prognosis of AIS. Recent studies have begun to explore the relationship between these biomarkers and symptomatic intracerebral hemorrhage (sICH) after EVT[18], as well as their potential for more accurate AIS diagnosis[19]. However, these studies have primarily focused on isolated aspects of the coagulation and fibrinolysis process, neglecting a holistic view. To our knowledge, there is a gap in the literature regarding the comprehensive exploration of the predictive effects of a range of coagulation function indicators on the prognosis of AIS patients post-EVT. Therefore, the objective of our study was to investigate the association between coagulation function (D-dimer, PT, INR, APTT, PTA, TT and FIB) and modern and long-term prognosis of patients with AIS-LVO after EVT.

## METHODS

### Study design and populations

This is a retrospective analyzing consecutive AIS patients with large vessel occlusion who underwent EVT from March 2016 to August 2023 in Figure 1. We included patients based on the following criteria: (1) patients who underwent thrombectomy employing second-generation stent-retriever devices or aspirator (Solitaire AB/FR, Covidien/ev3, Irvine, CA; Trevo Proview, Stryker, CA); (2) patients with occlusion of the large artery defined by digital subtraction angiography (DSA), including internal carotid artery (ICA), middle cerebral artery (MCA M1; M2), basilar artery(BA) and vertebral plus basilar artery (VPBA); (3) modified Rankin Scale (mRS) score before the index stroke ≤ 1; (4) EVT could be administered within 6 hours after symptom onset or within 6 – 24 hours after symptom onset with the presence of large ischemic mismatch/penumbra according to CT perfusion[20].

**Figure 1.**
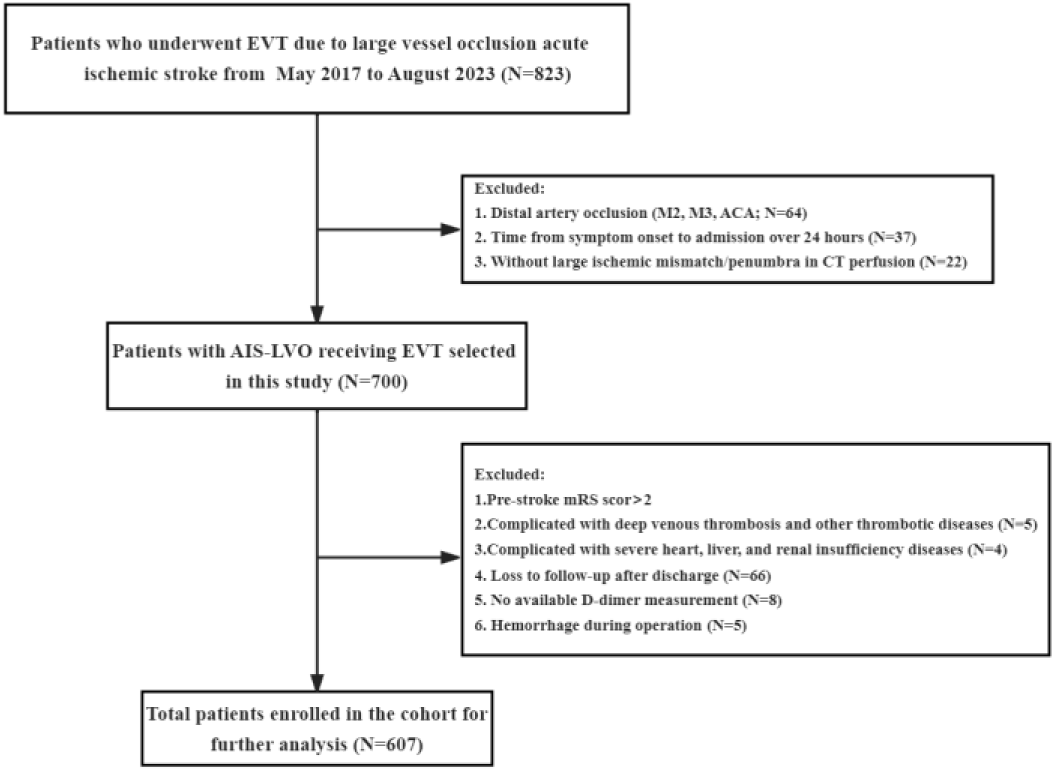
Flow chart of enrolled patients. EVT, endovascular thrombectomy; ICA, internal carotid artery; IVT, intravenous thrombolysis; ACA, anterior communicating artery; M2/M3, second/third segment of the middle cerebral artery; mRS, modified Rankin Scale; PE, pulmonary thromboembolism; sICH, symptomatic intracranial hemorrhage.

The exclusion criteria were as follows: (1) presence of intracranial hemorrhage identified on CT scan prior to EVT; (2) severe cardiac, hepatic, or renal insufficiency, or advanced diabetes mellitus with blood glucose levels exceeding 22 mmol/L; (3) coagulation disorders, such as platelet counts below 100×10^9^/L, or concurrent thrombotic conditions including but not limited to deep vein thrombosis, pulmonary embolism, and disseminated intravascular coagulation; (4) inability to obtain regular coagulation function tests within a 24-hour window preceding the operation or to compile complete medical records; (5) lack of follow-up imaging examinations or mRS scores after 3 months post-EVT. This rigorous selection process ensures a homogeneous study population that is representative of clinical practice while minimizing confounding factors that could influence the study outcomes.

### Data collection

Data for our study were extracted from the electronic medical records system, which is supported by the Stroke Center at the Department of the Second Affiliated Hospital, Zhejiang University. The patients included in our study underwent endovascular thrombectomy (EVT), encompassing a range of procedures such as thrombectomy with stent retrievers, thrombus aspiration, intracranial angioplasty, stent implantation, or any combination thereof, as determined by the treating surgeon. The occlusion sites of the arteries were identified through computed tomography angiography (CTA), magnetic resonance angiography (MRA), and/or cerebral digital subtraction angiography (DSA) reports, and included vessels such as the internal carotid artery (ICA), middle cerebral artery (MCA), basilar artery (BA), and vertebral artery (VERT).

The collected data were categorized into several domains, including baseline demographic data, medical history, stroke characteristics, procedural variables, treatment variables, and hospitalization complications, totaling 49 variables. Baseline characteristics encompassed age, sex, National Institutes of Health Stroke Scale (NIHSS) score, and modified Rankin Scale (mRS) score at admission, as well as comorbidities such as diabetes, hypertension, atrial fibrillation, hyperlipidemia, and a history of stroke, along with the use of antiplatelet and anticoagulation medications. Procedural time variables included metrics such as time from puncture to reperfusion (TPR), time from onset to groin puncture (OTP), time from onset to reperfusion (OTR), time from onset to admission (OTA), time from onset to imaging (OTI), and time from imaging to puncture (ITP). Additionally, we recorded the use of tissue plasminogen activator (t-PA), the number of retrieval attempts exceeding three, and rescue therapies including balloon angioplasty and stenting. Reperfusion status was assessed using the modified Thrombolysis in Cerebral Infarction (mTICI) scale by surgeons during the operation, with mTICI grades better than 2B indicating successful recanalization. Hospitalization complications such as hematoma transformation, symptomatic intracranial hemorrhage (sICH), respiratory failure, liver dysfunction, and pulmonary infection were also documented.

Blood samples were collected prior to the emergency EVT. Venous blood samples of 4-6 mL were obtained within 24 hours post-admission. In cases where blood tests were conducted multiple times within this timeframe, the initial value was utilized for analysis. Coagulation function indicators, including D-dimer, international normalized ratio (INR), prothrombin time (PT), prothrombin activity (PTA), activated partial thromboplastin time (APTT), thrombin time (TT), and fibrinogen (FIB), were measured using the Sysmex CA-7000 Automated Coagulation Analyzer (Sysmex, Kobe, Japan).[21].

### Follow-up and outcomes

The follow-up protocol was consistent with previously published literatures[22]. The primary outcome was good functional outcome at 90 days, defined as a modified Rankin Scale (mRS) score of 0–3[23]. The scores were collected by a stroke neurologist during routine follow-up visits at 90 days (±14) after stroke for the majority of patients through telephone discussion or clinical follow-up with patients or their families. As of complications, HT referred to the bleeding caused by the reperfusion of blood vessels in the ischemic area after acute cerebral infarction[24]. sICH was defined as any intracranial hemorrhage with an increase in the NIHSS score of≥4 from baseline) within 7days after EVT, according to the ECASS (European--Australasian Acute Stroke Study) II criteria[24].

### Statistical Analysis

Patients were dichotomized by favorable or unfavorable outcomes groups based on mRS score at 3 months after EVT were 0-3 or 4-6. Details on the missing data are shown in **online supplemental figure 1** without data missing more than 20%. For missing data sets, we use multiple interpolation to fill in the missing values. Characteristics are summarized as proportions for categorical variables and mean±SD or median (25-75th percentile) for quantitative variables, as appropriate. Two-sample t-tests or Mann-Whitney U test were used to compare the dichotomous in continuous quantitative variables, while χ2 tests or Fisher’s exact test were for categorical variables. Through ANOVA and univariate logistic regression, potential risk factors were filtrated to be associated with outcome of AIS patients (P<0.05), then these potential predictors were corrected by confounding factors which were filtrated from clinical characters through univariate logistic regression(P<0.05). Results were displayed as adjusted odds ratios (OR) combined with a 95% CI. The ROC was used to determine the cut-off value of predictors and their accuracy for the prediction of short-medium prognosis. Subgroup analysis was conducted to further analyze the different prognostic efficacy of coagulation function in certain subgroups. P values of < 0.05 were considered statistically significant. All reported P values were two-sided. All statistical analyses were performed using IBM SPSS Statistics for Windows version 26 and R language 4.0.5.

## Results

### Baseline characteristics of the cohort

The patient characteristics are detailed in **Table 1**. Our final cohort comprised 607 patients with a mean age of 69.10 ± 13.22 years. Of these, 259 (42.67%) were female, and 359 (59.14%) had received intravenous thrombolysis (IVT) prior to endovascular thrombectomy (EVT). The median National Institutes of Health Stroke Scale (NIHSS) score at admission was 15 (IQR, 11 −20), and the median modified Rankin Scale (mRS) score was 5 (IQR 3-5). On average, thrombectomy involved two pass attempts (IQR 1-3). At the conclusion of thrombectomy, 562 (92.59%) patients achieved successful recanalization, defined by a modified Thrombolysis in Cerebral Infarction (mTICI) score of 2B or 3.

**Table 1.** Baseline characteristics of patients with favorable or unfavorable outcomes after 3 months for endovascular treatment of acute large artery occlusive ischemic stroke.

| Variables | All patients(n = 607) | Good functional outcome<br>(n = 272) | Poor functional outcome<br>(n = 335) | P |
| --- | --- | --- | --- | --- |
| **Demographics characteristics** |  |  |  |  |
| Age, Mean ± SD | 69.10 ± 13.22 | 64.62 ± 13.70 | 72.74 ± 11.62 | <0.001 |
| BMI, Mean ± SD | 23.37 ± 3.44 | 23.68 ± 3.33 | 23.13 ± 3.51 | 0.054 |
| Female, n (%) | 259 (42.67) | 104 (38.24) | 155 (46.27) | 0.047 |
| Age>80, n (%) | 136 (22.41) | 29 (10.66) | 107 (31.94) | <0.001 |
| SBP, M (Q<sub>1</sub>, Q<sub>3</sub>) | 145.00 (128.00, 162.00) | 143.00 (124.75, 159.00) | 148.00 (129.00, 165.00) | 0.022 |
| CBP, M (Q<sub>1</sub>, Q<sub>3</sub>) | 81.00 (72.00, 91.00) | 80.50 (72.00, 90.00) | 82.00 (71.00, 92.00) | 0.73 |
| **Stroke characteristics** |  |  |  |  |
| NIHSS at admission, M (Q<sub>1</sub>, Q<sub>3</sub>) | 15.00 (11.00, 20.00) | 13.00 (9.00, 17.00) | 17.00 (12.00, 24.00) | <0.001 |
| NIHSS>14, n (%) | 401 (66.06) | 149 (54.78) | 252 (75.22) | <0.001 |
| mRS at admission, M (Q<sub>1</sub>, Q<sub>3</sub>) | 5.00 (3.00, 5.00) | 3.00 (2.00, 4.00) | 5.00 (5.00, 5.00) | <0.001 |
| TOAST classification, n (%) |  |  |  | 0.218 |
| LAA | 276 (46.85) | 140(52.63) | 136(42.11) |  |
| CE | 286 (48.56) | 110(41.35) | 176(54.49) |  |
| Others or undetermined | 18(2.97) | 6(2.21) | 12(3.58) |  |
| Site of occlusion, n (%) |  |  |  | <0.001 |
| ICA | 108 (17.79) | 43 (15.81) | 65 (19.40) |  |
| MCA | 319 (52.55) | 168 (61.76) | 151 (45.07) |  |
| ICA-MCA | 64 (10.54) | 23 (8.46) | 41 (12.24) |  |
| VERT/BA | 79 (13.01) | 19 (6.99) | 60 (17.91) |  |
| Others | 37 (6.10) | 19 (6.99) | 18 (5.37) |  |
| Multistage thrombus, n (%) | 106 (17.46) | 43 (15.81) | 63 (18.81) | 0.333 |
| **Procedural Variables** |  |  |  |  |
| OTA, M (Q<sub>1</sub>, Q<sub>3</sub>) | 266.00 (170.00, 374.00) | 256.00 (170.00, 373.00) | 270.50 (170.50, 375.75) | 0.967 |
| OTI, M (Q<sub>1</sub>, Q<sub>3</sub>) | 218.00 (115.75, 332.25) | 222.00 (124.00, 321.00) | 217.00 (111.50, 338.00) | 0.683 |
| ITP, M (Q<sub>1</sub>, Q<sub>3</sub>) | 104.00 (76.00, 138.00) | 99.00 (76.00, 130.50) | 107.27 (76.00, 141.00) | 0.142 |
| PTR, M (Q<sub>1</sub>, Q<sub>3</sub>) | 70.00 (45.00, 105.72) | 63.00 (43.60, 96.00) | 75.00 (50.00, 113.50) | 0.005 |
| OTR, M (Q<sub>1</sub>, Q<sub>3</sub>) | 400.00 (300.00, 521.00) | 391.00 (296.50, 509.00) | 413.50 (305.00, 529.50) | 0.087 |
| OTP, M (Q<sub>1</sub>, Q<sub>3</sub>) | 325.00 (230.00, 431.50) | 320.00 (230.44, 419.01) | 330.50 (230.00, 440.00) | 0.431 |
| Numbers of pass attempts, M (Q<sub>1</sub>, Q<sub>3</sub>) | 2.00 (1.00, 3.00) | 2.00 (1.00, 3.00) | 2.00 (1.00, 3.00) | 0.007 |
| Numbers of pass attempts >3, n (%) | 98 (16.14) | 34 (12.50) | 64 (19.10) | 0.028 |
| Successful recanalization (mTICI≥2B/3), n(%) | 562 (92.59) | 259 (95.22) | 303 (90.45) | 0.026 |
| **Treatment Variable** |  |  |  |  |
| #Rescue therapy | 105 (17.30) | 50 (18.38) | 55 (16.42) | 0.525 |
| Tirofiban treatment, n (%) | 105 (17.30) | 56 (20.59) | 49 (14.63) | 0.053 |
| Distal escape, n (%) | 42 (6.92) | 20 (7.35) | 22 (6.57) | 0.704 |
| IVT, n (%) | 359 (59.14) | 165 (60.66) | 194 (57.91) | 0.493 |
| General Anesthesia, n (%) | 311 (51.24) | 126 (46.32) | 185 (55.22) | 0.029 |
| **Medical History** |  |  |  |  |
| CIA, n (%) | 134 (22.08) | 53 (19.49) | 81 (24.18) | 0.166 |
| Hyperlipemia, n (%) | 22 (3.62) | 10 (3.68) | 12 (3.58) | 0.951 |
| Hypertension, n (%) | 391 (64.42) | 152 (55.88) | 239 (71.34) | <0.001 |
| CHD, n (%) | 75 (12.36) | 24 (8.82) | 51 (15.22) | 0.017 |
| Diabetes, n (%) | 114 (18.78) | 32 (11.76) | 82 (24.48) | <0.001 |
| Atrial fibrillation, n (%) | 231 (38.06) | 86 (31.62) | 145 (43.28) | 0.003 |
| Pre-antiplatelet, n (%) | 72 (11.86) | 36 (13.24) | 36 (10.75) | 0.346 |
| Pre-anticoagulation, n (%) | 84 (13.84) | 35 (12.87) | 49 (14.63) | 0.532 |
| Operation history, n (%) | 205 (33.77) | 83 (30.51) | 122 (36.42) | 0.126 |
| **Hospitalization complications** |  |  |  |  |
| HT, n (%) | 205 (33.77) | 51 (18.75) | 154 (45.97) | <0.001 |
| siCH, n (%) | 133 (21.91) | 4 (1.47) | 129 (38.51) | <0.001 |
| Pulmonary infection, n (%) | 282 (46.46) | 103 (37.87) | 179 (53.43) | <0.001 |
| Liver dysfunction, n (%) | 74 (12.19) | 32 (11.76) | 42 (12.54) | 0.772 |
| Respiratory failure, n (%) | 53 (8.73) | 7 (2.57) | 46 (13.73) | <0.001 |
| **Coagulation function** |  |  |  |  |
| D-dimer | 970.00 (505.00, 2205.00) | 665.00 (380.00, 1472.50) | 1290.00 (630.00, 2690.00) | <0.001 |
| INR, M (Q<sub>1</sub>, Q<sub>3</sub>) | 1.04 (0.98, 1.11) | 1.02 (0.96, 1.08) | 1.05 (0.98, 1.13) | 0.002 |
| APTT, M (Q<sub>1</sub>, Q<sub>3</sub>) | 33.90 (31.25, 37.15) | 33.90 (31.20, 36.82) | 33.80 (31.30, 37.45) | 0.731 |
| TT, M (Q<sub>1</sub>, Q<sub>3</sub>) | 17.20 (16.30, 18.20) | 17.10 (16.20, 18.20) | 17.20 (16.40, 18.20) | 0.527 |
| PT, M (Q<sub>1</sub>, Q<sub>3</sub>) | 13.40 (12.90, 14.20) | 13.30 (12.80, 13.90) | 13.50 (13.00, 14.30) | 0.002 |
| PTA, M (Q<sub>1</sub>, Q<sub>3</sub>) | 94.00 (84.00, 104.00) | 96.00 (87.00, 106.00) | 93.00 (82.00, 103.00) | 0.004 |
| FIB, M (Q<sub>1</sub>, Q<sub>3</sub>) | 3.07 (2.63, 3.67) | 2.96 (2.53, 3.49) | 3.18 (2.67, 3.82) | 0.002 |
| Abbreviation: CHD, Coronary heart disease; MRS, Modified Rankin Scale; ICA, Internal carotid artery; MCA, Middle cerebral artery, NIHSS, National Institutes of Health Stroke Scale; TPR, time from puncture to reperfusion; OTP, time from onset to groin puncture; OTR, time from onset to reperfusion; OTA, Time from Onset to admission; OTI, Time from Onset to image; ITP, Time from image to puncture; LAA, large artery atherosclerosis; CE, cardioembolism; IVT, intravenous thrombolysis; mTICI, modified Thrombolysis in Cerebral Infarction; HT, Hemorrhagic transformation; siCH, symptomatic intracranial hemorrhage, INR, international normalized ratio; APTT, activated partial thromboplastin time; TT, thrombin time; PT, prothrombin time; PTA, prothrombin time activity; FIB, fibrinogen. |  |  |  |  |
| Qualitative variable are n (%), mean ± SD, median (interquartile range). * p<0.05; ** p<0.001. |  |  |  |  |
| # Rescue therapy included balloon angioplasty and permanent stenting. |  |  |  |  |

Within the cohort, 335 (55.19%) AIS-LVO patients exhibited an unfavorable outcome, characterized by an inability to walk independently three months later. Those with an unfavorable prognosis were older on average (72.74 vs. 64.62 years), more frequently female (46.27% vs. 38.24%), had higher systolic blood pressure (SBP) at emergency presentation (median 148 vs. 143 mmHg), and experienced longer times from puncture to recanalization (median 75 vs. 63 minutes). They also presented with higher NIHSS scores (median 17 vs. 13) and mRS scores (median 5 vs. 3). The prevalence of comorbidities was higher among patients with poor outcomes, including hypertension (13.73% vs. 2.57%), atrial fibrillation (43.28% vs. 31.62%), coronary heart disease (CHD) (15.22% vs. 8.82%), and diabetes (24.48% vs. 11.76%). Hospitalization complications were also more prevalent in this group, with higher rates of hematoma transformation (HT) (45.97% vs. 18.75%), sICH (38.51% vs. 1.47%), pulmonary infection (53.43% vs. 37.87%), and respiratory failure (13.73% vs. 2.57%). In term of coagulation function, patients with unfavorable prognosis had higher level of D-dimer (median 1290 vs 665μg/L), INR (median 1.05 vs 1.02), PT (median 13.5vs 13.3 seconds) and PFI (median 3.18vs 2.96 g/L) but lower PTA (median 93% vs 96%) compared to those with favorable prognosis after EVT (**Table 1)**. Other baseline characteristics and coagulation function indicators, like stroke etiology or APTT didn’t have significant differences between two different groups (P>0.05).

### Adjusted odds ratio of D-dimer, INR, PT and PFI with poor prognosis at 3 months after EVT

After adjusting for variables found to be significant in univariate logistic analysis, we ensured that the tolerance of the variables included in the multivariate logistic analysis was greater than 0.1, and the variance inflation factor (VIF) was significantly less than 10 for all predictors. This indicated the absence of multicollinearity within our models. Elevated levels of D-dimer (aOR = 1.01; 95% 1.01 to 1.01) and decreased prothrombin activity (PTA) (aOR = 0.99; 95% CI 0.98 to 0.99) were significantly associated with a poor outcome in patients with AIS-LVO following EVT (**Table 2**). Additionally, an increase in the international normalized ratio (INR) showed a trend towards an increased risk of a poor prognosis, although this did not reach statistical significance (aOR = 2.14; 95% CI 0.93 to 4.96, P = 0.074). In the adjusted clinical variables, age above 80 years (aOR = 3.89; 95% CI 2.25 to 6.73), National Institutes of Health Stroke Scale (NIHSS) score above 14 (aOR = 2.28; 95% CI 1.50 to 3.45), diabetes (aOR = 2.45; 95% CI 1.43 to 4.20), basilar artery (BA) occlusion (aOR = 3.08; 95% CI 1.43 to 6.61), and atrial fibrillation (aOR = 1.53; 95% CI 1.01 to 2.34) were independently associated with a poor prognosis (**Online Supplemental Table 1**).

**Table 2.** Unadjust and adjusted ORs of the association of coagulation function indicators with poor outcome at 3 months after endovascular treatment.

| Variables | Univariate |  | Multivariate |  |  |
| --- | --- | --- | --- | --- | --- |
| | OR (95% CI) | $P$ | $a\beta$ | aOR (95% CI) | $aP$ |
| Ddimer | 1.01 (1.01 ~ 1.01) | 0.006 | 0.01 | 1.01 (1.01 ~ 1.01) | 0.011 |
| INR | 1.98 (1.01 ~ 3.86) | 0.045 | 0.76 | 2.14 (0.93 ~ 4.96) | 0.074 |
| PT | 1.00 (0.98 ~ 1.02) | 0.803 |  | — |  |
| PTA | 0.99 (0.98 ~ 0.99) | 0.009 | -0.01 | 0.99 (0.98 ~ 0.99) | 0.017 |
| FIB | 1.29 (1.09 ~ 1.53) | 0.003 | 0.15 | 1.16 (0.96 ~ 1.41) | 0.131 |

Adjust procedural variables incorporate male, age higher than 80 years, successful recanalization (mTICI=2B or 3), general anesthesia, NIHSS higher than 14, hypertension, CHD, diabetes, site of occlusion, atrial fibrillation, pass attempts over 3 times, body fat index, PTR and baseline SBP.
OR: Odds Ratio, CI: Confidence Interval. \* $p < 0.05$ ; \*\* $p < 0.001$ .

### Diagnostic efficiency of D-dimer and PTA with operating characteristic curve

Based on the receiver operating characteristic curve (ROC) in **Figure 2**, the optimal cut-off value of D-dimer as a predictor for predicting poor outcome was 715 mg/L, which showed a sensitivity of 71.3% and a specificity of 54.1% with the AUC of 0.647 (95% CI 0.603 to 0.692). In the same way, the optimal cut-off value of PTA was 85.5% with a sensitivity of 79.3% and a specificity of 33.4%, and the AUC of 0.578 (95% CI, 0.522 to 0.613). Through cut-off value, we further divided patients into the low D-dimer group (D-dimer <715 mg/L) and the elevated D-dimer group (D-dimer≥715 mg/L). Similarly, we also divided patients into low PTA group (PTA< 85.5%) and the elevated PTA group (PTA≥85.5%). The clinical characteristics of patients with normal and elevated group are shown in **online supplemental table 2**. What’s more, after adjustment for significant variables in univariate analysis, male (acOR=0.61; 95%CI 0.39 to 0.96), basilar artery occlusion (acOR=0.32; 95%CI 0.12 to 0.84), baseline mRS score (acOR=1.24; 95%CI 1.06 to 1.46) were significantly associated with baseline elevated D-dimer levels **(online supplemental table 3)**. For emergency PTA, CE stroke etiology (acOR=0.43; 95%CI 0.25 to 0.77), operation history (acOR=0.4; 95%CI 0.24 to 0.67), history of taking anticoagulant drugs (acOR=0.26; 95%CI 0.14 to 0.5) were significantly associated with baseline elevated PTA after correcting for confounding variables **(online supplemental table 4).**

**Figure 2.**
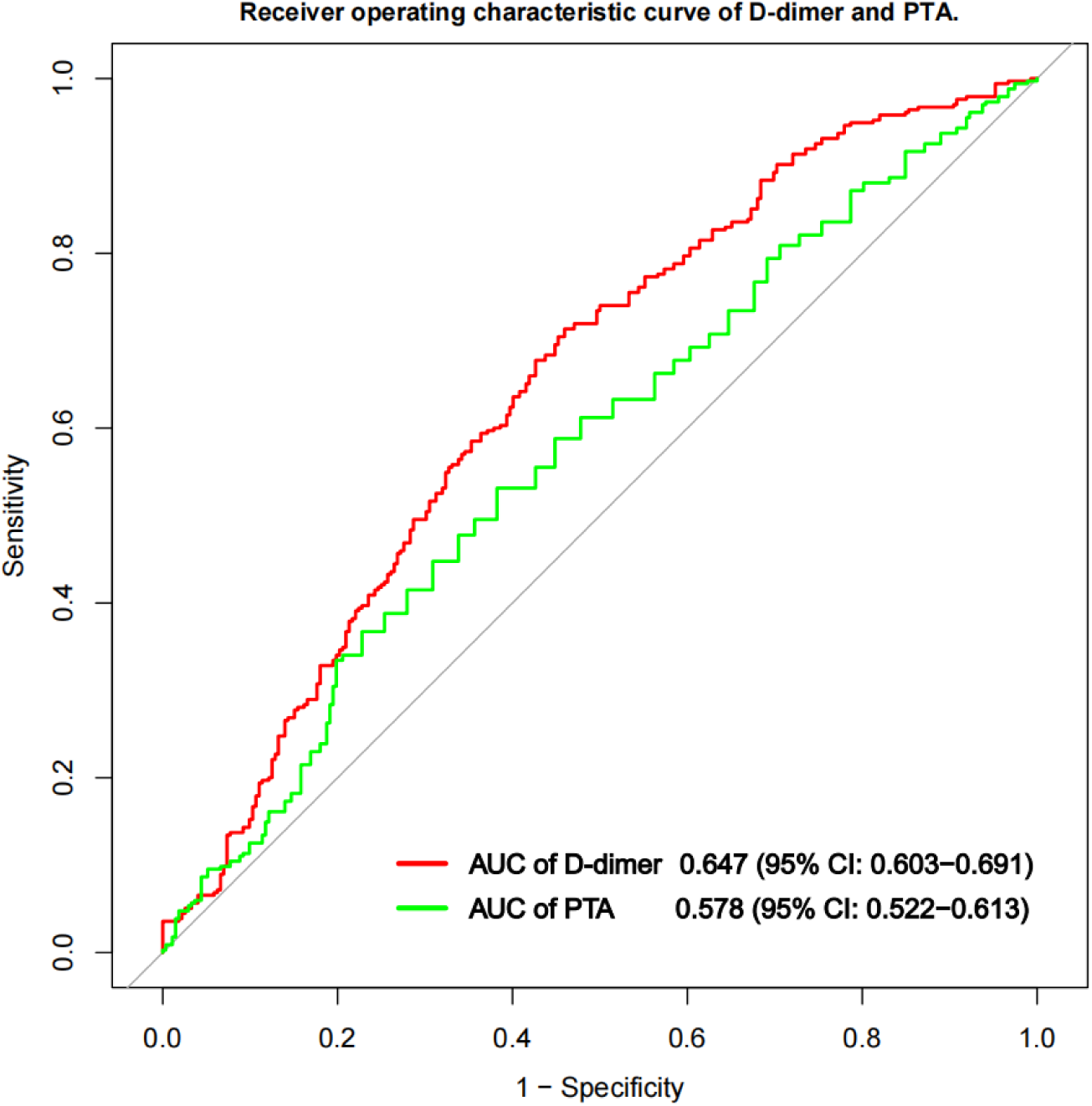
Receiver operating characteristic (ROC) curve was used to evaluate the predictive ability of plasma D-dimer and NLR levels for 3-months unfavorable prognosis of AIS patients after EVT (area under the curve=0.647 and 0.578 respectively).

We further explore the association between elevated D-dimer and PTA group and poor prognosis of AIS in **Table 3**. After adjusting for other covariates, elevated D-dimer was the independent predictor of poor prognosis, with 2.51 times higher in the risk of poor outcome in elevated D-dimer (≥715 mg/L) than normal D-dimer group (aOR=2.51, 95CI 1.59 to 3.97). But the elevated PTA(≥85.5%) was independently and inversely linked to poor prognosis, and the risk of poor prognosis was 0.39 times in the elevated PTA group than the normal group (aOR=0.39, 95%CI 0.18 to 0.87). In addition, elevated D-dimer and low PTA had synergistic effect, presenting higher association with poor outcome of patients with AIS-LVO (aOR=4.55, 95%CI 2.63 to 7.69).

**Table 3.**
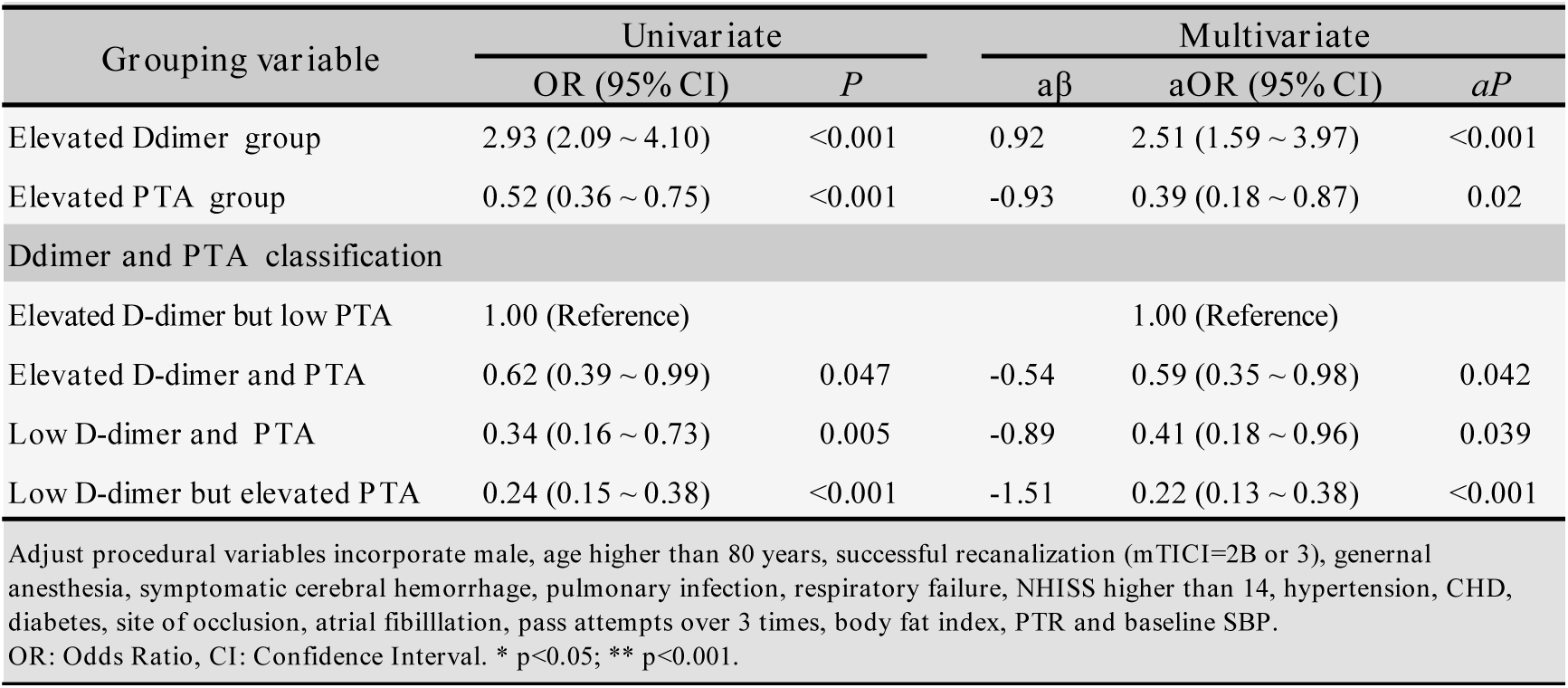
Unadjust and adjusted ORs of the association of coagulation function indicators with poor outcome at 3 months after endovascular treatment.

### Subgroup analysis of elevated D-dimer and PTA group

Subgroup variables were selected from positive clinical risk factors after multivariate correction **(Figure 3)**. D-dimer exhibits a stable predictive efficacy on prognosis of AIS across different subgroups. There is no significant interaction with nine stratification factors (Interaction P>0.05). Subgroup analysis showed that except in a subgroup of patients who did not achieve complete recanalization (mTICI<2B/3), complicated with diabetes and not the MCA occlusive stroke, high D-dimer group were significantly associated with a worse prognosis after EVT for patients with AIS (P<0.05). Conversely, for PTA, only in male, age lower than 80 years, successful recanalization, NHISS higher than 14, without diabetes, with atrial fibrillation, MCA occlusion, and pass attempt less than 3 times subgroup, elevated PTA rate was significantly associated with a better prognosis for patients with AIS. Patients with failed recanalization, complicated with diabetes and not the MCA occlusive, these three subgroups affect the predictive efficacy of both D-dimer and PTA for clinical outcome, which deserve more attentions in clinical practice.

**Figure 3A.**
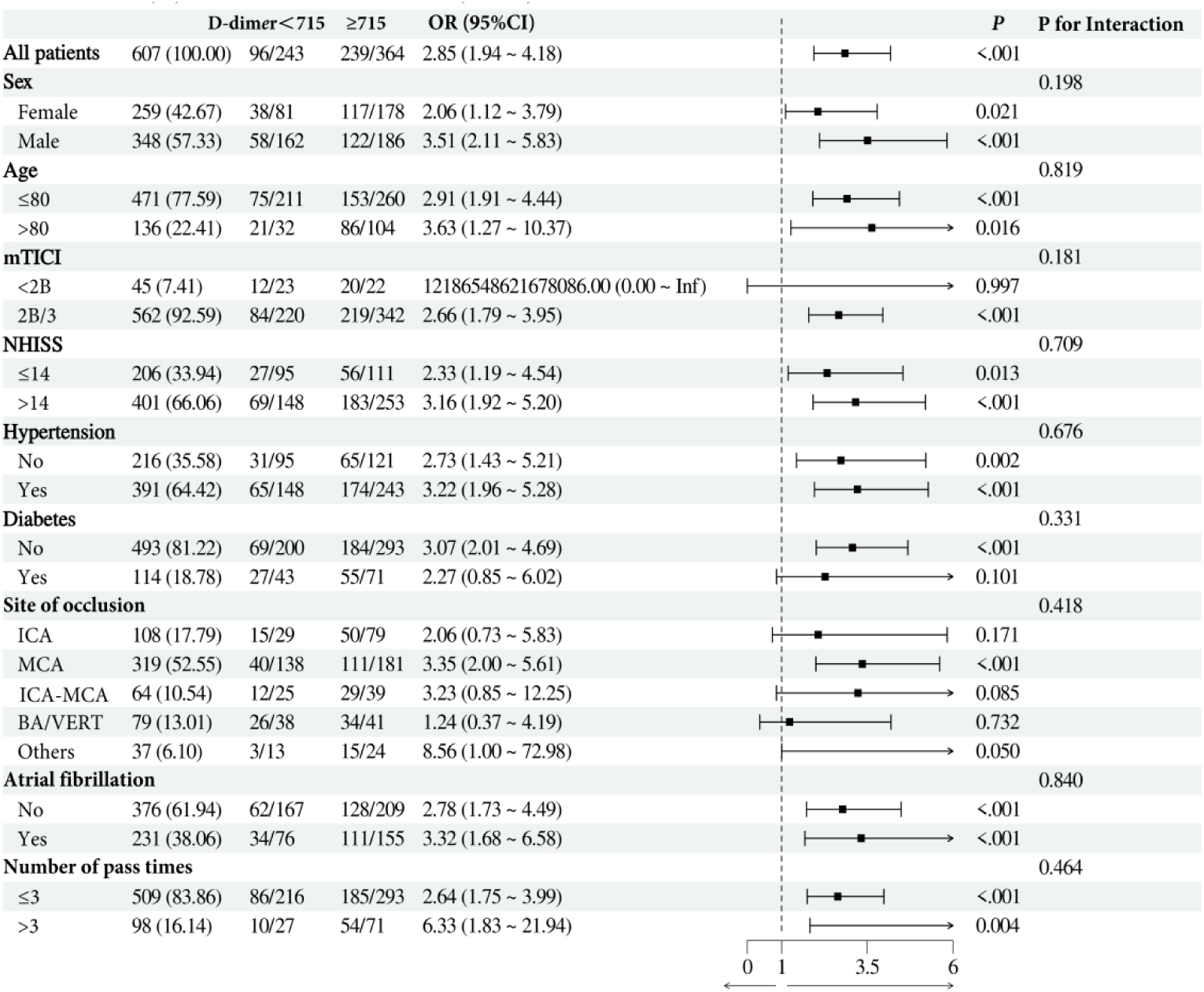
Subgroup analysis of unfavorable prognosis in elevated D-dimer group. There are no interaction effects in subgroup of sex, age, mTICI, NHISS, hypertension, diabetes, site of occlusions, atrial fibrillation and number of pass times.

**Figure 3B.**
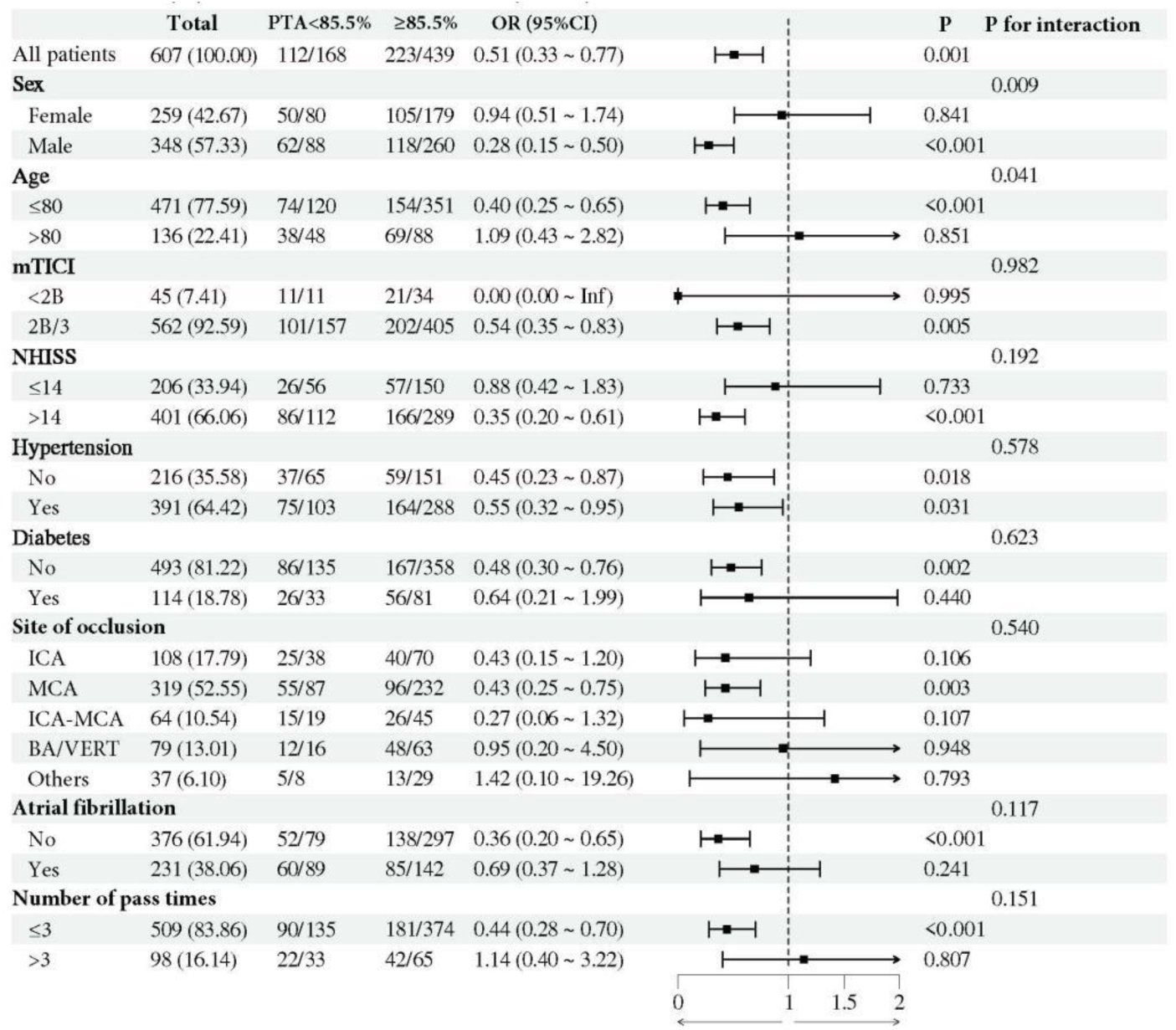
Subgroup analysis of unfavorable prognosis in elevated PTA group. Elevated PTA presented significant interaction effect with subgroup of age. There are no interaction effects in subgroup of sex, mTICI scores, NHISS scores, hypertension, diabetes, site of occlusions, atrial fibrillation and number of pass times.

## Discussion

In clinical guideline, the key to the treatment of AIS is to rapid restoration of blood supply to ischemic brain tissue as soon as possible and salvage ischemic penumbra[25]. Nowadays, with more updated retriever devices and various kinds of thrombectomy techniques having been stabilized, the successful recanalization proportions of EVT to between 83% and 92%[26]. In the latest clinical trials, results showed even higher recanalization rates with up to 95% of patients achieving successful recanalization[18]. However, the rate of favorable clinical outcomes still wanders around 50% even lower[27]. Therefore, it is urgent and important to explore which factors cause and influence the mismatch between technical success rates and clinical outcomes, which will be beneficial to guide clinical treatment decisions and make personalized treatment plan according to different conditions of patients and their families. As a routine clinical preoperative examination, we systemically investigated the associations between coagulation function profile (PT, INR, PTA, TT, APTT and FIB) and unfavorable prognosis of patients with AIS at 3 months after EVT. After adjusting for other covariates, D-dimer was the independent predictor of poor prognosis, with 2.51 times higher in the risk of poor outcome in elevated D-dimer group than low D-dimer group (aOR=2.51, 95CI 1.59 to 3.97). But in high level of PTA was independently and inversely linked to poor prognosis, and the risk of poor prognosis was 0.39 times in the elevated PTA group than in the normal group (aOR=0.39, 95%CI 0.18 to 0.87). In addition, elevated D-dimer and low PTA had synergistic effect, presenting higher association with poor outcome of patients with AIS-LVO (aOR=4.55, 95%CI 2.63 to 7.69). The increase of INR had a tendency to increase the risk of poor prognosis (P = 0.074), and no correction between PT, APTT and FIB and poor outcome was found. Subgroup analysis showed that three subgroups, incorporating failed recanalization, complicated with diabetes and not the MCA occlusive affected the predictive efficacy of both D-dimer and PTA for clinical outcome, which deserve more attentions in clinical practice.

As a soluble fibrin degradation product that results from ordered breakdown of thrombi by the fibrinolytic system, D-dimer exhibit a close connection with AIS. Numerous studies have shown that D-dimer serves as a valuable marker of activation of coagulation and fibrinolysis[28]. The increase of D-dimer concentration is a sensitive marker of fibrinolysis secondary to acute thrombosis, which is closely related to the occurrence, development and prognosis of stroke[29]. In ischemic stroke, the baseline elevated D-dimer levels may reflect ongoing microthrombosis in the target vessel occlusion territory, thus causing extensive activation of the coagulation system, subsequent hyperfibrinolysis and coagulopathic disturbances[18]. Han et al.[30] reported that AIS patients are at increased risk of developing DVT following EVT particularly if they have undergone prolonged thrombectomy procedures and exhibit high plasma levels of D-dimer (aOR 1.35, 95%CI 1.15 to 1.59). Wang Q et al.[31] demonstrated that D-dimer levels before reperfusion were independent predictors for 3-months unfavorable outcome in AIS patients undergoing reperfusion therapy (no matter intravenous thrombolysis and/or endovascular). In atrial fibrillation patients, D-dimer to fibrinogen ratio independently predicts early neurological deterioration (END) in ischemic stroke with adjusted odds ratio (aOR= 2.14, 95%CI 1.24 to 3.6)[32]. More importantly, a latest study demonstrate that elevated admission D-dimer level was an independent predictor of sICH in patients with AIS after thrombectomy (aOR=2.45, 95%CI 1.75 to 3.43). Based on the ROC, the D-dimer as a predictor for predicting sICH, presented with a specificity of 86.2%, and an area under the curve of 0.774[18]. Besides, it have been demonstrated that HT, especially sICH, plays a counteract role in functional outcomes of AIS following EVT[27]. So, D-dimer levels may indirectly influence patients’ prognosis of 3-months by influencing the occurrence of procedure complications.

PTA is an important indicator reflecting the synthesis function of coagulation factors in the liver, which was usually done by a functional measurement of prothrombin in plasma, reflecting the actual functional level of prothrombin in blood clotting[33]. When liver hepatocytes are seriously damaged, the reduction of coagulation factor synthesis PTA will decrease. Therefore, the level of PTA had been reported that can reflect the degree of hepatocyte damage, which is of great value for the condition evaluation and prognosis of patients with liver failure[34]. No previous studies have reported the prognostic effect of PTA in stroke patients. Our study found that the level of PTA was a protective factor for ischemic stroke prognosis. The risk of poor prognosis in AIS-LVO patients after EVT with elevated PTA (>85.5%) was 0.39 times compared with that in patients with low PTA group (aOR=0.39, 95%CI 0.18 to 0.87), showing liver function may indirectly affect the prognosis of stroke patients. As the indicator of the liver’s ability to produce coagulation factors, effective hepatoprotective treatment and enhancement of the liver’s capacity to produce coagulation factors may become a new method to assist in the functional recovery of stroke patients in the clinical practices.

PT is a commonly index to assess the status of exogenous coagulation function, which measures the time of activity of clotting factors in the blood, specifically the time of activation of clotting factor I (prothrombin) into thrombin in vitro[21]. INR is closely related to PT, which is a standardized method of comparing a patient’s PT results to an internationally standardized sample to eliminate variability between different laboratories, different reagents, and different test equipment[33]. In the present study, we found no association between PT/INR and poor prognosis of AIS patients after EVT, which was consistent with previous studies[35,36]. As calculated by PT, INR directly reflected the status of blood coagulation, and can be elevated by anticoagulation treatment[33]. In our cohort, proportion of patients with atrial fibrillation was 38.06%, which may affect hemodynamics and coagulation function to affect INR rate. Therefore, the extrinsic pathway of coagulation may not have a vital role in affecting prognosis, but therapeutic interventions that may increase level of PT and INR in acute ischemic stroke patients should be selected carefully. During blood clotting, FIB is activated together with prothrombin and converted into fibrin, forming a web of fibers that eventually leads to the formation of blood clots. Thrombin can cleave FIB into fibrin to form blood clots and exert impacts on fibrinolysis in different directions[37]. Furthermore, the structure of a fibrin clot was found related to functional outcomes in ischemic stroke patients[38]. In our research, we found there are no association between FIB and poor prognosis. In our study, we only investigated the emergency level of FIB of patients with AIS-LVO and found no association. However, as a dynamic, The FIB level will fluctuate in different time periods with the progression of ischemic region and blood reperfusion. Hence, the dynamic change of FIB and its role in the prognosis of AIS deserves further study.

In addition to D-dimer and NLR, we also found some other factors, such as higher age, sex, use of tilofiban, and mRS at admission are also independently associated with prognosis of AIS. Previous studies have shown that the prognosis of elderly patients after mechanical thrombectomy (MT) is relatively poor. The reason is that most elderly patients take anticoagulants and antiplatelet drugs for a long time due to other diseases, which may lead to secondary cerebral hemorrhage[39]. In addition, due to atherosclerosis and intravascular plaque formation in elderly patients, the operation of MT is more difficult. In addition, the probability of complications is higher than that of younger patients, and the tolerance to surgery is worse than that of younger patients[40]. Studies have reported variable results for stroke case fatality by sex, with many providing little evidence of a difference, some showing higher case fatality, and others reporting lower case fatality for women. Overall, baseline differences in age, stroke characteristics, and cerebrovascular risk factors account for much of the observed sex differences in case fatality. Age-adjusted mortality rates are commonly reported, but because sex differences are strongly modified by age, they can mask the complex relation of sex differences across age groups and can hide the burden of stroke mortality for elderly women[41].

There are still some limitations in our study: Firstly, it is based on single-center data, which may not capture the full clinical profile and endovascular treatment trends for AIS-LVO patients. Secondly, as a retrospective study, it is susceptible to selection bias, and therefore, the results should be interpreted cautiously. Thirdly, the time span of our cohort is long from 2016 to 2023, which may cause deviation due to the progress of materials and the maturity of surgical techniques to improve the treatment effect and prognosis. Finally, we did not track and analyze serial D-dimer levels post-thrombectomy, a parameter that could be key to understanding patient outcomes. Further studies are warranted to determine whether dynamic D-dimer and PTA changes are also associated with the moderate and long-term prognosis in AIS-LVO. Exploring the pathophysiological mechanism of association between D-dimer and PTA levels may beneficial to explain the cause of poor functional prognosis of patients with AIS after thrombectomy.

## Conclusions

In conclusion, our study systemically explored preoperative coagulation function and revealed that D-dimer and PTA levels were independent predictors of 3-month prognosis after thrombectomy in patients with AIS-LVO. Besides, INR showed a tendency to increase the risk of poor prognosis but no significance was found. Elevated D-dimer group (>715μg/L) was a risk factor for poor prognosis but the elevated PTA (≥85.5%) was a protective factor. In addition, elevated D-dimer and low PTA had synergistic effect, presenting higher adjusted odds ratio with prognosis after 3 months. After subgroup analysis, Patients with failed recanalization, complicated with diabetes and not the MCA occlusive, these three factors affect the predictive efficacy of both D-dimer and PTA for clinical outcome, which deserve more attentions in clinical practice. Given that quantitative measurement of D-dimer and PTA levels are easily obtainable in clinical practice, their use as a blood biomarker predicting prognosis after EVT may add early prognostic information and facilitate perioperative management.

## Data Availability

Funder requirements are not applicable, but data will be available within the article or in an appropriate repository at time of publication.

## Declarations

### Ethical Approval and Consent to participate

This large retrospective study was approved by human Research Ethics Committee, the Second Affiliated Hospital of Zhejiang University School of Medicine. Without causing any potential harm to the patient, the patient’s informed consent is exempt.

### Consent for publication

Not applicable.

### Availability of data and materials

Data and material are not publicly available, but can be requested.

### Authors’ contributions

Conception and design: S.J. and P.Z.

Acquisition of data: C.Q. and J.Y. and X.L.

Analysis and interpretation of data: L.X.

Drafting of the article: S. J. and P.Z.

Critically revising the article: F.B. and Z.T.

Reviewed submitted version of manuscript: all authors.

Approved the final version of the manuscript on behalf of all authors: J.L. and J.X.

Administrative/technical/material support: X.C. and F.B.

Study supervision: J.L.

## Acknowledgments

This work was financially supported by the Natural Science Foundation of Zhejiang Province (LQ21H090008) and Zhejiang medical and health science and technology project (2021434317). We also thank Lingzhi Yu for her comments on the manuscript.

